# Population-weighted greenspace exposure tied to lower COVID-19 mortality rates: A nationwide dose-response study in the United States

**DOI:** 10.1101/2022.05.24.22275549

**Authors:** Yuwen Yang, Yi Lu, Bin Jiang

## Abstract

The COVID-19 outbreak has caused enormous deaths and profound social and economic disruption globally. Accumulating evidence suggests exposure to greenspace may reduce the risk of COVID-19 mortality. Greenspace exposure enhances immune functioning, reduces inflammation, and replenishes gut microbiota may protect against the risk of mortality among those with COVID-19. However, previous studies often fail to distinguish the health effect of different types of greenspace, explore the dose-response association and optimal buffer distance, and consider the spatial dynamics of population distribution and geographic locations of greenspace.

This study examined the associations among ratio of different types of greenspaces, population-weighted exposure to different types of greenspaces, and COVID-19 mortality rates using a negative binomial generalized linear mixed effects model across 3,025 counties in the United States, adjusted for socioeconomic, demographic, pre-existing chronic disease, policy and regulation, behavioral, and environmental factors. The population-weighted measure gave proportionally greater weight to greenspace near areas of higher population density.

Exposure to forest and pasture was negatively associated with COVID-19 mortality rates, while developed open space has insignificant or positive associations with mortality rates. *Forest outside park* has the largest effect size across all buffer distances, followed by *forest inside park*. The optimal exposure buffer distance is 1km for *forest outside park*, with 1 unit of increase in exposure associated with a 9.9% decrease in mortality rates (95% confidence interval: 6.9% -12.8%). The optimal exposure buffer distance of *forest inside park* is 400m, with 1 unit of increase in exposure, associated with a 4.7% decrease in mortality rates (95% confidence interval: 2.4% - 6.9%).

Greenspaces, especially nearby forest, may be effective at lowering the mortality risk of COVID-19 patients. Our findings suggest that policymakers and planners should prioritize forestry within walking distance of residential clusters to mitigate mortality rates during current and future respiratory pandemics.

## 1. Introduction

Since its outbreak in 2019, COVID-19 has spread rapidly throughout the world, leading to numerous infections and deaths. In the United States, COVID-19 is largely responsible for the substantial 17.7% increase in total deaths from 2019 to 2020 and became the third leading cause of death following heart disease and cancer (Ahmad, Cisewski, Miniño, & Anderson, 2021). By the end of 2020, COVID-19 deaths were estimated at 348,600; by February 2020, they had reached over 933,000 (Johns Hopkins University & Medicine, 2021; Viglione, 2020). In the US, COVID- 19 was estimated to reduce life expectancy by 1.13 years annually (Andrasfay & Goldman, 2021).

The COVID-19 pandemic also overwhelmed healthcare systems and caused substantial economic loss. COVID-19 infection and deaths cast escalating pressure on testing capacities and hospitalizations in the U.S. (Dyer, 2020; Miller, Becker, Grenfell, & Metcalf, 2020). Critically ill COVID-19 patients faced shortages in intensive care units (ICUs) compounded by other critical health conditions (Halpern & Tan, 2020). The cumulative economic costs of the COVID-19 pandemic due to premature deaths, unemployment, and business revenue decline was estimated to be US$1.4 trillion GDP by 2030 (Cutler & Summers, 2020; Chen et al. 2021).

Accumulating evidence suggests links between both nature and the built environment and COVID-19 mortality rates. Exposure to air pollution (Ali & Islam, 2020; Konstantinoudis et al., 2021; Liang et al., 2020), crowded housing (Brandén et al., 2020; Hu, Roberts, Azevedo, & Milner, 2021; van Ingen et al., 2021), and lower average temperature (Ma et al., 2020; Perone, 2021) were found to increase COVID-19 deaths. The relationship between greenspace and COVID-19 mortality rates has received far less attention (Jiang, Yang, et al., 2021; Klompmaker et al., 2021; Lu, Chen, et al., 2021), despite the numerous salutary effects of nature exposure on human health.

### 1.1 How might exposure to greenspace alleviate COVID-19 mortality rates?

An overwhelming amount of research has shown that exposure to greenspace can improve both physical and mental health (Jiang, Chang, & Sullivan, 2014; Lu, Chen, et al., 2021). Particularly, studies have shown that contact with greenspace boosts our defense capacity against viruses by increasing Natural Killer (NK) and T cells and cytotoxic activities (Liisa Andersen, Sus Sola Sola Corazon, & Ulrika Karlsson Karlsson Stigsdotter, 2021; Li, 2010; Roviello, Gilhen-Baker, Vicidomini, & Roviello, 2021), reducing inflammation (Kuo, 2015; Ribeiro, Tavares, Guttentag, & Barros, 2019), and replenishing gut microbiota (Parajuli, 2019; Parajuli et al., 2020; Marja I. Roslund et al., 2020). Hospitalized COVID-19 patients with severe or fatal cases have consistently shown immune interference (e.g., lower NK and T cell count, exaggerated cytotoxic activities) (Castelli, Cimini, & Ferri, 2020; Girija, Shankar, & Larsson, 2020; Qin et al., 2020), hyper- inflammation or ‘cytokine storm’(e.g., delayed but elevated of pro-inflammatory cytokines) (Paranjpe et al., 2020; Potempa, Rajab, Hart, Bordon, & Fernandez-Botran, 2020; Yang et al., 2020), and decreased gut microbiota diversity (Dhar & Mohanty, 2020) compared to non-critically ill patients. Thus, contact with nature has the potential to mitigate severe COVID-19 prognosis and deaths.

### 1.2 A critical gap: The relationship between different types of greenspace and COVID-19 mortality rate

Several studies have shown a significant association between greenness and COVID-19 mortality rates in the U.S. (Klompmaker et al., 2021; Lee et al., 2021; Russette et al., 2021; Spotswood et al., 2021). These studies define greenspace as the total area of vegetation within a boundary (e.g., Normalized Difference Vegetation Index and Leaf Area Index).They did not distinguish between open space, forest, grassland/herbaceous, and hay/pasture, nor did they consider the impacts of greenspace proximity and recreational function provision on COVID-19 health outcomes. While previous evidence suggests different types of greenspace does not have same impacts on health outcomes (Akpinar, Barbosa-Leiker, and Brooks 2016; Ekkel & de Vries 2017; Kim & Miller 2019; Johnson et al. 2020; Ma et al. 2022). For instance, greenspace such as greenness and park had negative association with COVID-19 infection (Russette et al. 2021; Spotswood et al. 2021; Wang et al. 2021; Johnson et al. 2020), but park mobility and green space with better accessibility were positively associated with COVID-19 transmission (Pan, Bardhan, and Jin 2021; DePhillipo et al., 2021). We still do not know whether and to what extent different types of greenspace can influence the COVID-19 mortality rates.

Second, existing studies estimate the amount of greenness in a county but ignore the spatial distribution of greenspace in relation to population (Klompmaker et al., 2021; Russette et al., 2021). Despite being widely used and effective, the accuracy of ‘greenness’ metric can be greatly improved by considering spatial relations between location of greenspace and population distribution (Ben et al., 2019). Further, one study used deciles of ‘greenness’ to assess the dose-response association (Russette et al., 2021). Though, the dose-response associations for different types of greenspace within various buffer distances are unclear. Many previous studies suggested that distance matters for greenspace’s impact on health outcomes, and health effect might drop after a threshold distance (Coombes, Jones, and Hillsdon 2010; Grahn and Stigsdotter 2003; Nielsen and Hansen 2007). Still, we do not know whether the association of nearby greenspace on mortality risk is significantly stronger than distant ones. We do not know which distances are optimal. Without addressing these critical gaps, policymakers and urban planners are unable to develop evidence-based urban greening solutions and policy to promote public health for current and future pandemics.

### 1.3 Research questions

In this study, we investigated the associations among the ratio of six types of greenspace, population-weighted exposure to six types of greenspace at different buffer distances, and full-year COVID-19 mortality rates after controlling for potential confounding covariates. We seek to answer the following three questions: 1) What are the associations between the ratio of six types of greenspace and COVID-19 mortality rates after controlling for confounding variables? 2) What are the associations between population-weighted exposure to six types of greenspace and COVID-19 mortality rates within various buffer distances after controlling for confounding variables? (3) Which exposure distances of the significant types of greenspace have the strongest association with COVID-19 mortality rates?

## 2. Methods

We combined COVID-19 mortality data, sociodemographic characteristics, healthcare and testing data, pre-existing chronic disease data, policy and regulation data, behavior data, and environmental factors from diverse sources for 3,025 counties. The greenspace exposures were calculated in GEE. We first used a negative binomial generalized linear mixed effects model to evaluate the association between the ratio of six types of greenspace and COVID-19 mortality rates in the U.S. from January 22 to December 31, 2020, adjusted for socioeconomic, demographic, pre-existing chronic disease, policy and regulation, behavioral, and environmental factors. Then, we examined the associations between the population-weighted exposure to greenspace at varying distances within 4km and COVID-19 mortality rates, adjusted for confounders.

### 2.1 Data

#### 2.1.1 COVID-19 mortality data

COVID-19 mortality data are publicly available at the US Centers for Disease Control and Prevention (CDC) and State government websites (Kolak et al., 2021). We define the COVID-19 mortality rates as the cumulative number of COVID-19 deaths per 100,000 people for each of 3,025 counties from January 22, 2020 to December 31, 2020 (Fig. 1). We limit our research period at the end of 2020 to avoid the possible confounding effect from large-scale vaccination, which will have significantly impact on mortality rates (see Supplementary Table 1 for descriptive COVID-19 mortality data).

**Fig. 1.**
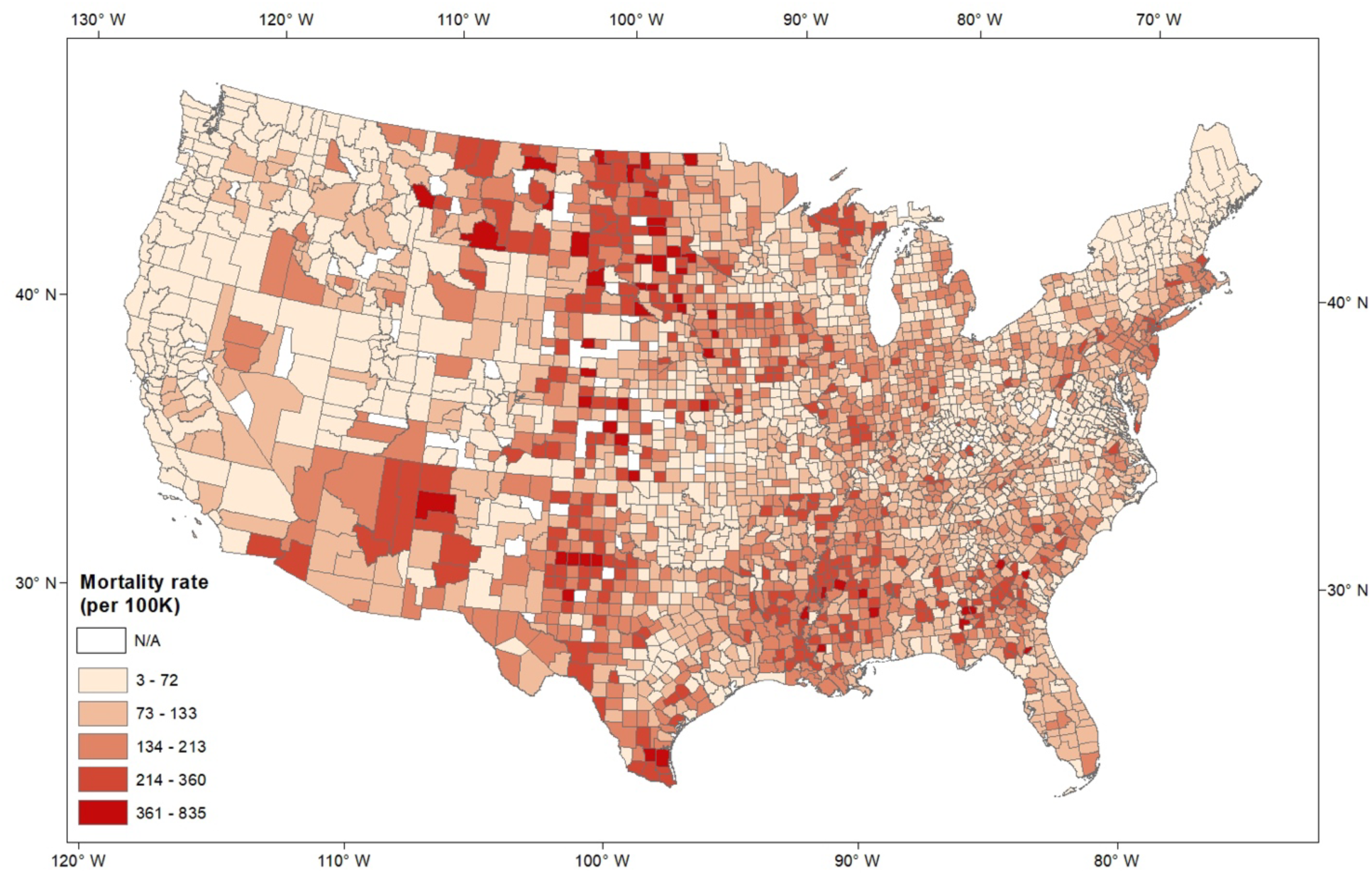
County-level COVID-19 deaths per 100,000 population in the United States. (from Jan 18 to Dec 30, 2020).

#### 2.1.2 Greenspace exposure data

We assessed greenspace exposure using two metrics. First, we quantified the ratios of six types of greenspaces within a county, which measures the area of each greenspace over the county area. The ratio of *forest inside park, forest outside park*, grassland/herbaceous, hay/pasture, *open space inside park*, and *open space outside park* were calculated use the National Land Cover Database 2016 (NLCD, 2016) (Fig. 2). We distinguished open space and forest within park from open space and forest outside park use the boundary derived from the USA parks from Esri (Esri, 2021).

**Fig. 2.**
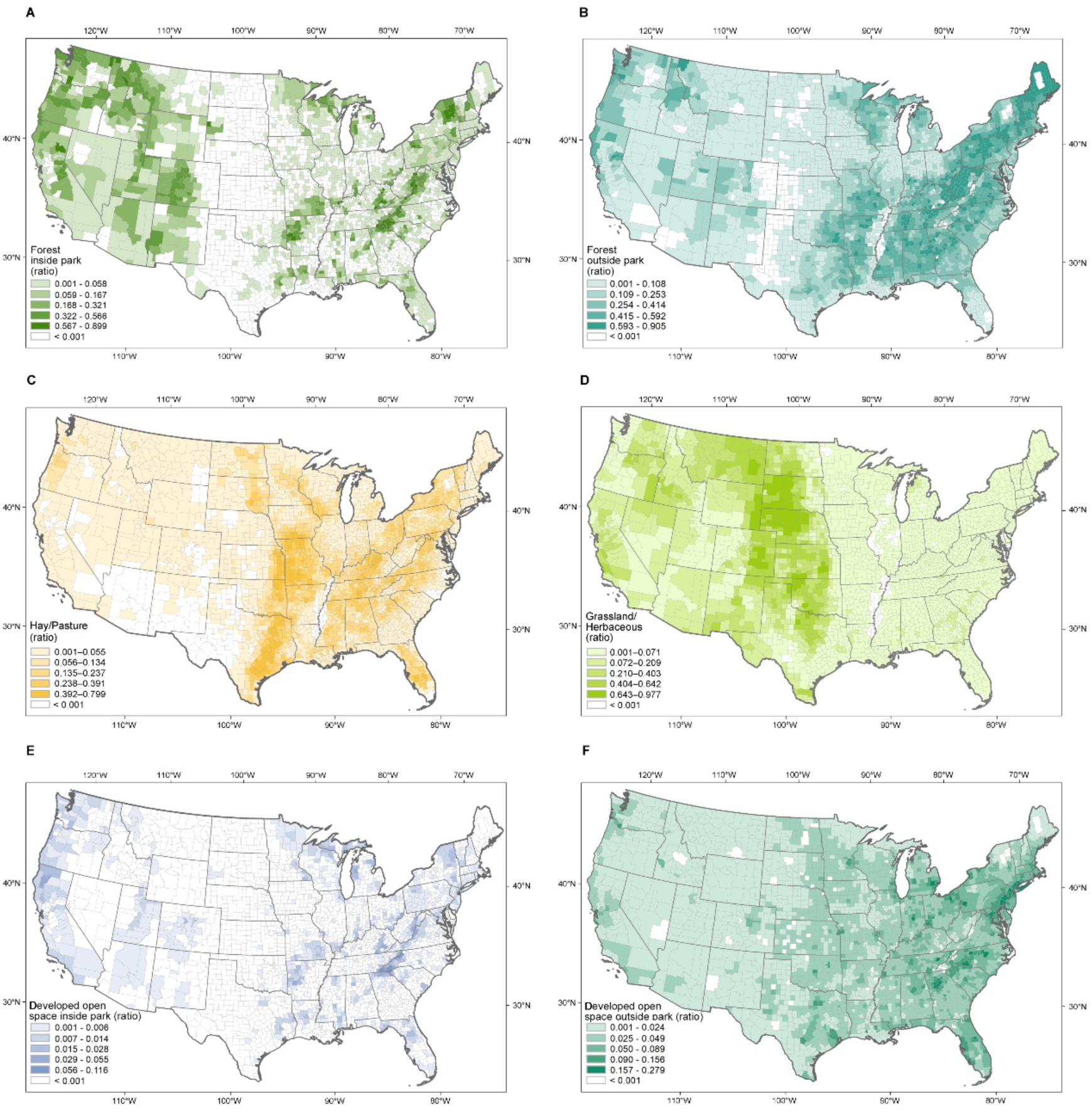
Ratio of greenspace at the county level in the United States. (**A**) forest inside park (**B**) forest outside park; (**C**) hay/pasture; (**D**) grassland/herbaceous; (**E**) developed open space inside park; (**F**) developed open space outside park.

Second, we quantified the population-weighted exposure to greenspaces within different distances from human settlements using two datasets: the National Land Cover Datasets in 2016 (Yang et al., 2018) and the 2020 WorldPop Global Project Population Data (Sorichetta et al., 2015). The 30-meter resolution NLCD 2016 Landsat imagery was re-projected to match the 100- meter spatial resolution of WorldPop Dataset. We estimated population-weighted greenspace exposures within 4km in GEE (Gorelick et al., 2017), because past studies suggest few walking activities occur beyond 4km (Yang & Diez-Roux, 2012). These measures considered population spatial distributions and gave proportionally greater weight to greenspace near areas of higher population density, which previous studies fail to address (Chen et al., 2018). Considering population distribution in greenspace exposure measurement can reduce bias caused by a mismatch between population and greenspaces within an area. The buffer interval is set as 200m within 2km and 500m between 2km to 4m. The population-weighted exposure to greenspaces with varying buffer sizes in each county is defined by Equation 1 (Chen et al., 2018),

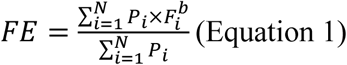

where *P*_*i*_ represents the population of the *i*^th^ grid, 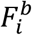 represents the land cover of the *i*^th^ grid at a buffer size of *b* meters, *N* denotes the total number of grids for a given county, and *FE* is the estimated greenspace exposure level for the given county (see Supplementary Table 2 for descriptive greenspace exposure data).

#### 2.1.3. Predictors of COVID-19 mortality

We considered a number of predictors of COVID-19 mortality as potential covariates in analyses. Many studies have found sociodemographic, chronic disease, behavioral, healthcare, and environmental factors linked to COVID-19 mortality. The county-level sociodemographic, healthcare, and testing data from the US Census Bureau (US Census Bureau, 2019) and US COVID Atlas of the Center for Spatial Data Science (Kolak et al., 2021). These variables included population density, the ratio of female household, non-Hispanic black, white, and Hispanic, the proportion of residents older than 65, median household income, Gini index, poverty rate, median housing value, unemployment rate, Gini index, the ratio of residents without a high school diploma and without a college degree, percent without health insurance coverage, and COVID-19 testing rates.

The pre-existing chronic diseases were shown to affect COVID-19 mortality risk, included rates of hypertension, heart failure, stroke mortality, diabetes, and obesity (CDC, 2021; Prevention., 2022). The policy and regulation factors included stay-at-home order intensity, public mask mandates, and bar and restaurant closing and reopening orders (Chernozhukov et al., 2021; VoPham et al., 2020). The behavior risk factors included the proportion of current smokers and the proportion of essential workers, the proportion of workers who commuted to work by public transportation, walking, and private cars, and the proportion of leisure-time physical inactivity, median max-distance traveled, and foot traffic to different out-of-home activities. The environmental risk factors included particulate matter (e.g., PM_2.5_ and PM_10_), temperature, relative humidity, precipitation, wind speed, and transportation density. All descriptive statistics of covariates and data sources are available in Supplementary Table 1.

### 2.2 Statistical analysis

We used a negative binomial generalized linear mixed effects model to evaluate the associations between the ratios of six types of greenspace and COVID-19 mortality rates, and state was used in analyses as a random effect to account for state-level variability and non-independence in our data. The analyses were adjusted for a range of covariates. We applied restricted maximum likelihood (REML) with a negative binomial link function. The negative binomial mixed effect model and state as random effect accounts for our over-dispersed count data and partially accounts for the presence of spatial autocorrelation. The variance inflation factor (VIF) test was used to identify multi-collinearity between the independent variables. Variables with a VIF ≥ 4 were excluded from our models (O’Brien, 2007).

To identify the optimal exposure distance for significant greenspaces, we used a negative binomial generalized linear mixed effects model to evaluate associations between population-weighted exposures to six types of greenspace with COVID-19 mortality rates. The analyses use the same sets of covariates as previous analysis and state was used as random effect. All explanatory variables were centered and scaled.

We used Moran’s I test to assess spatial autocorrelation of COVID-19 mortality residuals. We confirmed the presence of spatial autocorrelation with Moran’s *I*=0.21, *p* < 0.0001. The Moran’s I value equal to 0 indicates a lack of spatial autocorrelation, and positive values indicate clustering of similar values. The analyses were performed in R v.4.1.2 (Team, 2015), and Moran’ I test was performed using the package ‘spdep’ (Bivand & Wong, 2018). The negative binomial mixed effects models were performed using the package lme4 (Bates, Mächler, Bolker, & Walker, 2014).

### 2.3 Model validation

The negative binomial generalized linear mixed effects model provides an appropriate error structure for the overdispersed COVID-19 mortality count data. Due to the presence of spatial autocorrelation (Moran’s *I*=0.21, *p* < 0.0001), we built additional spatial autoregressive models (SAR) to validate the results of the negative binomial mixed effects model. The queen’s criteria were used to build the neighbors matrix. We used the Akaike information criterion (AIC) value to compare the spatial error model, spatial lag model, and spatial Durbin model. The spatial error model has the lowest AIC values, which suggests that spatial dependence occurs in the error term. The model validation confirms the negative associations between *forest inside park, forest outside park*, pasture, and COVID-19 mortality rates (See results of SAR models in Supplementary Table 5). Given the structure of our data, model coefficients and the magnitude of effects, we chose to interpret our result using the negative binomial generalized linear mixed effects model.

## 3. Results

### 3.1 Associations among ratio of six types of greenspaces and COVID-19 mortality rates

We found *forest inside park* and *forest outside park* are significantly negatively associated with COVID-19 mortality rates (*p* < 0.0001); *open space outside park* is significantly positively associated with COVID-19 mortality rates (*p* < 0.01); grassland/herbaceous, hay/pasture and *open space inside park* are not significantly associated with COVID-19 mortality rates, after controlling for all covariates (Fig. 3). Among the six types of greenspace, *forest outside park* has the greatest effect size (*β* = -0.097), which is slightly larger than that of *forest inside park* (*β* = -0.082). We found 1 unit of increase in *forest outside park* is associated with a 9.2% decrease in COVID-19 mortality rates (MRR 95% CI: 6.3 - 12.1%), whereas 1 unit of increase in *forest inside park* is associated with a 7.8% decrease (MRR 95% CI: 4.3 - 11.2%). In contrast, 1 unit of increase in *developed open space outside park* is associated with a 5.8% increase in COVID-19 mortality rates (MRR 95% CI: 2.3 - 9.5%) (Table 1).

**Table 1.**
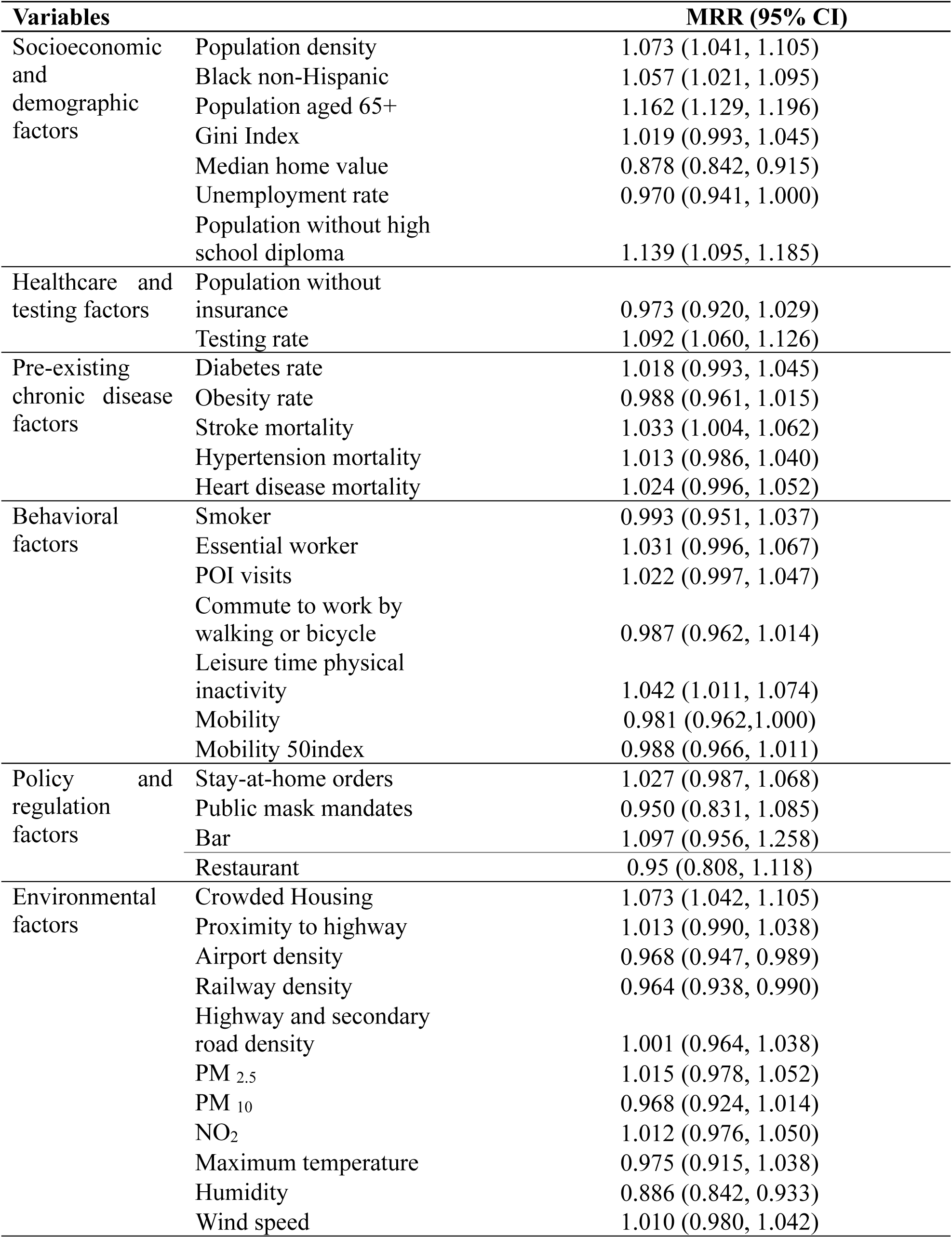
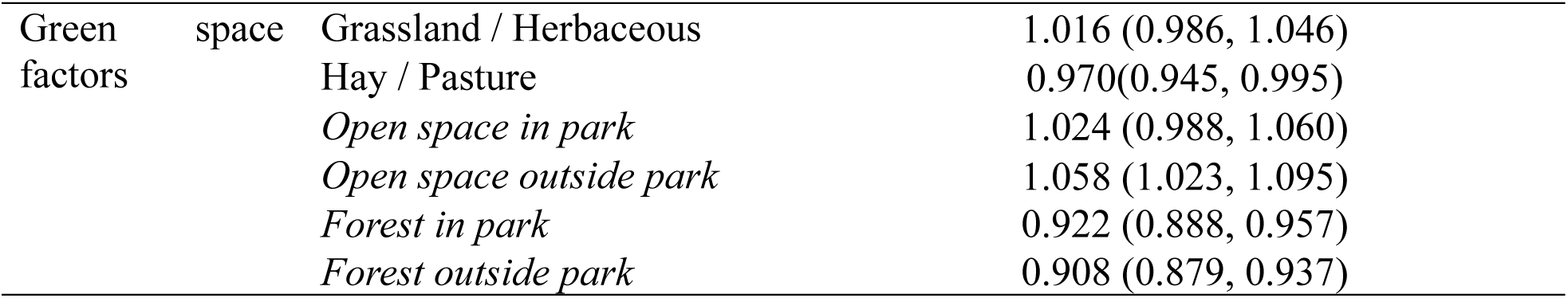
Mortality rate ratios (MRR), 95% confidence intervals (CI) of all variables in the model for COVID-19 mortality rates.

**Fig. 3.**
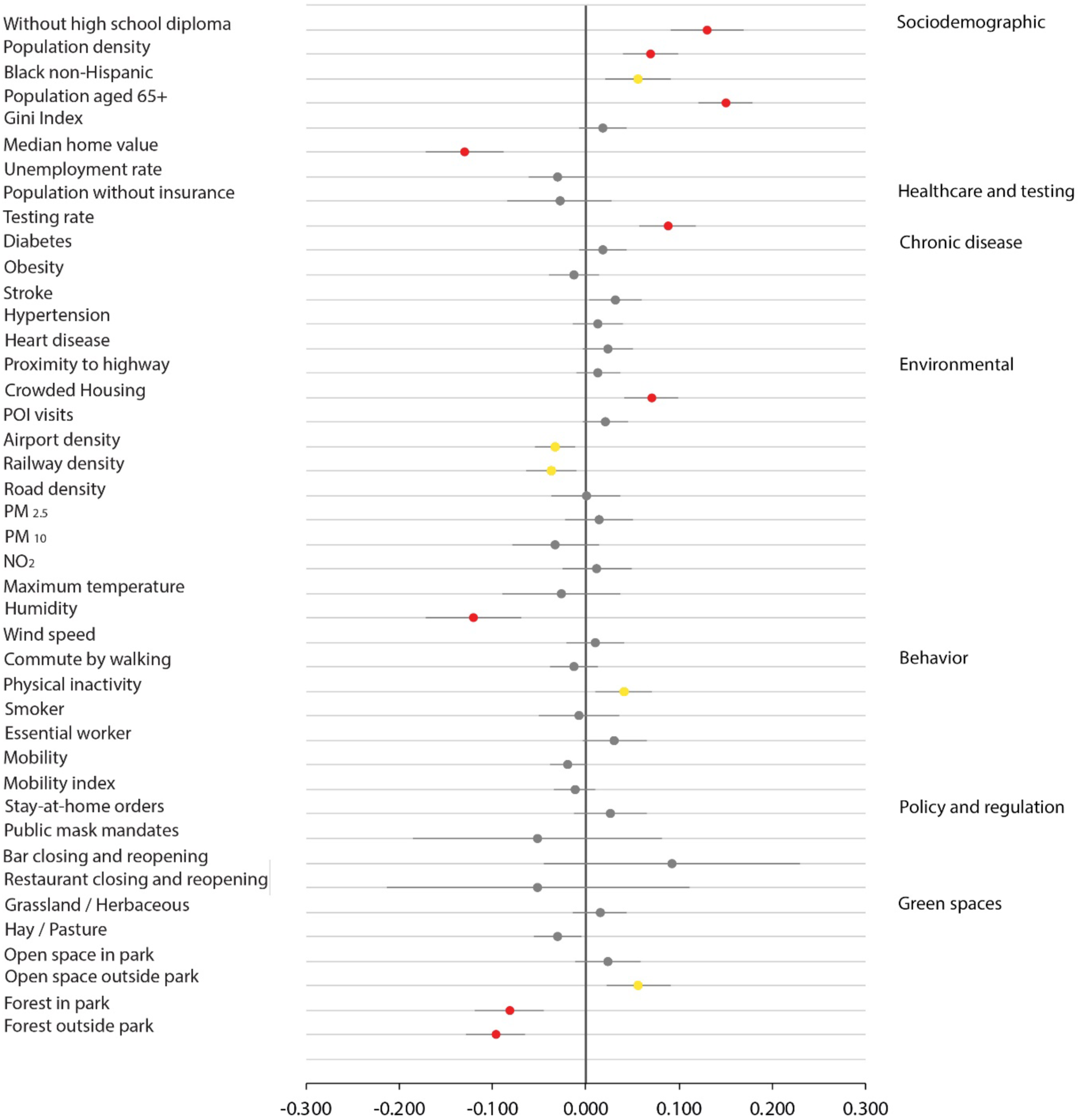
Exposure to forest is associated with lower COVID-19 mortality rates adjusted for covariates. Coefficient values represent effect sizes for the associations between mortality rates of COVID-19 (cases per 100,000 people) and ratio of grassland/herbaceous, hay/pasture, *open space in park, open space outside park, forest inside park, forest outside park*, and all covariates. Coefficient values are represented as dots, bars represent 95% CI, and significant variables are shown in color: grey = *p* ≥ 0.01; yellow = *p* < 0.01; red = *p* < 0.0001.

### 3.2 Associations of population-weighted exposures to greenspace with COVID-19 mortality rates at various buffer distances

We also found population-weighted exposure to *forest inside park, forest outside park* and pasture are significantly and negatively associated with COVID-19 mortality rates. Population-weighted exposure to *forest inside park* is significantly and negatively associated with mortality rates between 100 to 400m and 1,800m to 4km, and the distance for largest effect size is 4km (*β* = -0.050). The effect size increases as buffer distance gets larger, though the increase remains limited (400m *β*= -0.048 vs 4km *β* = -0.050, 4% of increase) (Fig. 4). With 1 unit of increase in *forest exposure in park* at 4km, there is a 4.9% decrease in COVID-19 mortality rates (MRR 95% CI: 2.7-7.0%) (Table 2).

**Fig. 4.**
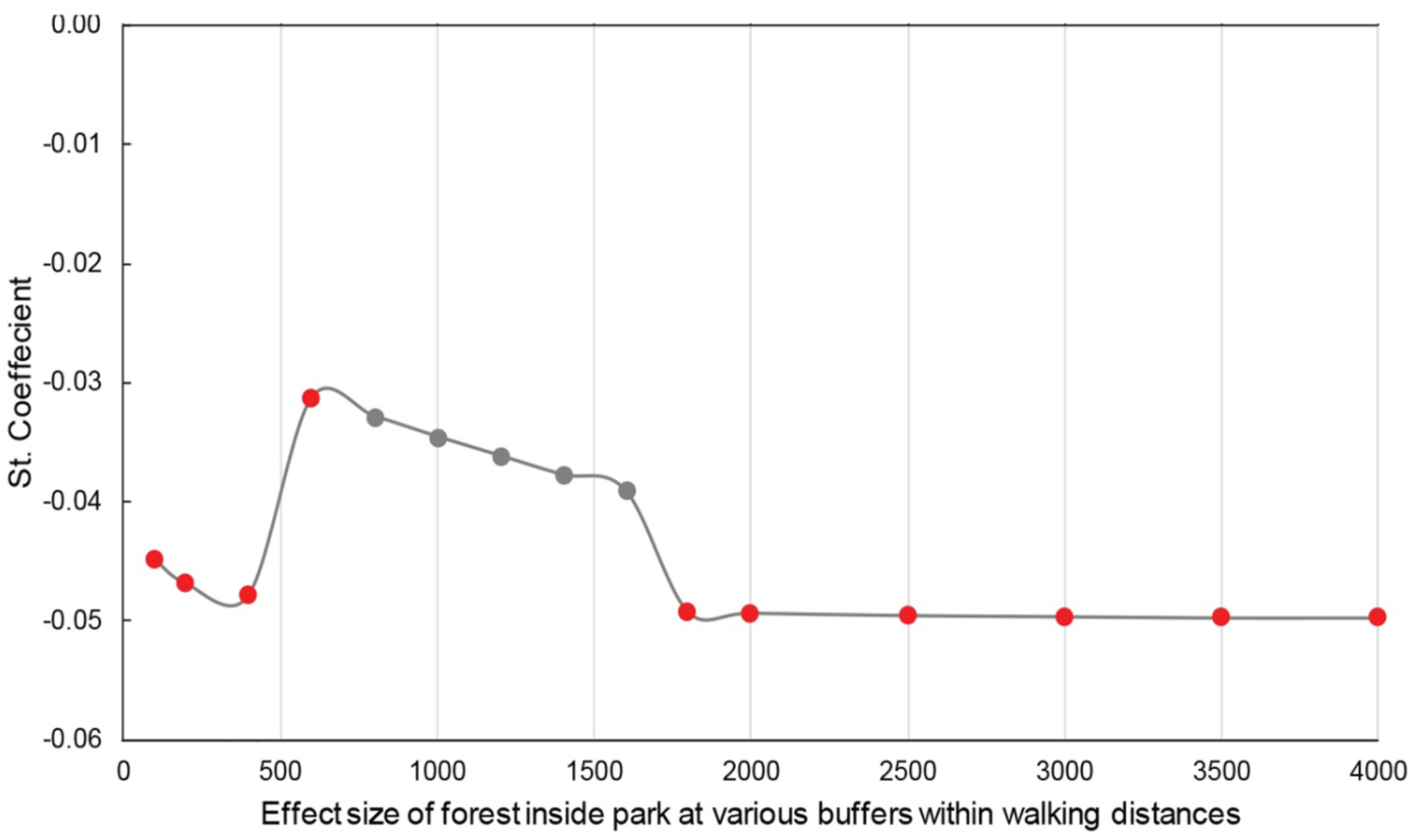
The effect size of population-weighted exposure to *forest inside park* within 4km on COVID-19 mortality rates. Coefficient values represent effect sizes from a negative binomial mixed effects model for the relationship between mortality of COVID-19 mortality rates (death count per 100,000 people) and population-weighted exposure to *forest inside park*. Coefficient values are represented as dots, grey = *p* > 0.05; red = *p* < 0.0001.

The population-weighted to *forest outside park* is consistently and significantly negatively associated with COVID-19 mortality rates across all buffer distances, and the greatest reduction occurs at 1km (*β* = -0.104). The effect size increases as the buffer increases from 100m to 1km and decrease beyond 1km (Fig. 5). We found a 9.9% decrease in COVID-19 mortality rates per unit increase in *forest exposure outside park* at 1km (MRR 95% CI: 6.9–12.8%) (Table 2).

**Table 2.** Mortality Rate Ratio in models of population-weighted exposure to greenspaces with COVID-19 mortality rates at optimal distances.

| Variables | COVID-19 MRR (95% CI) |
| --- | --- |
| *Forest inside park 400m* | 0.953 (0.931, 0.976) |
| *Forest inside park 4km* | 0.951 (0.930, 0.973) |
| *Forest outside park 1km* | 0.901 (0.872, 0.931) |
| Hay/pasture 4km | 0.965 (0.941, 0.990) |

**Fig. 5.**
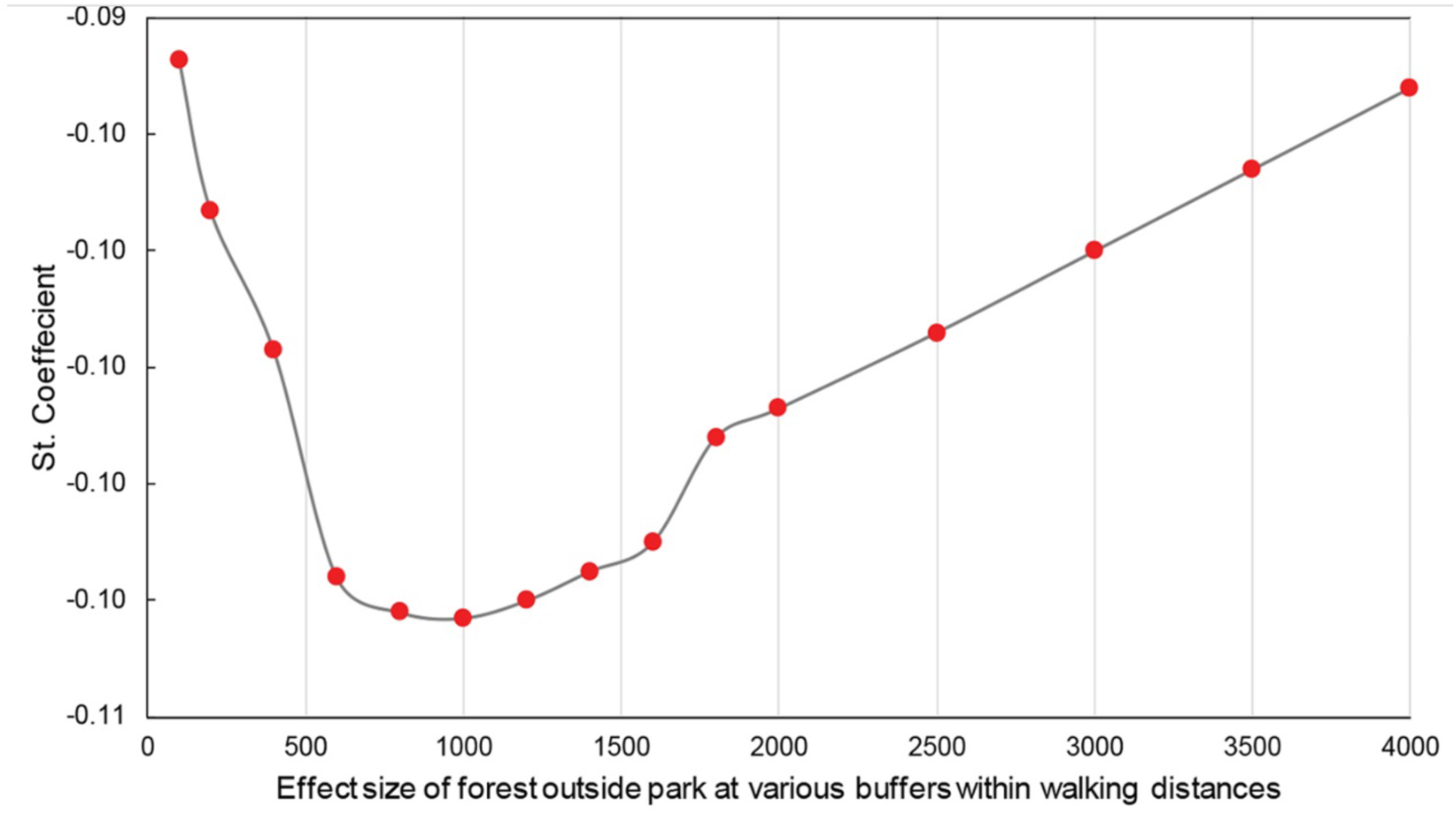
The effect size of population weighted exposure to *forest outside park* within 4km on COVID-19 mortality rates. Coefficient values represent effect sizes from a negative binomial mixed effects model for the relationship between mortality of COVID-19 mortality rates (death count per 100,000 people) and population-weighted exposure to *forest outside park*. Coefficient values are represented as dots, red = *p* < 0.0001.

The population-weighted exposure to *pasture* is significantly negatively associated with mortality rates from 2,500m to 4km with increasing effect size, and reaches optimal effect at 4km (*β* = -0.036). With 1 unit of increase in pasture exposure at 4km, the associated mortality rates decrease by 3.5% (MRR 95% CI: 5.9 - 10.0%) (Table 2).

## 4. Discussion

We found exposure to forest and pasture to be negatively associated with COVID-19 mortality rates, while exposure to developed open space has mixed association with mortality rates. Exposure to *forest outside park* has the largest effect size on reduced COVID-19 mortality rates across all buffer distances, followed by *forest inside park*. Further, the effect size of exposure to *forest outside park* increases until the distance reaches 1km, then declines beyond 1km. The effect size of population-weighted exposure to *forest inside park* increase with larger buffer size and is greatest at 4km, though similar to that at 400m.

While this cross-sectional study cannot infer any causal relationships, previous findings suggest multiple mechanisms that might explain the observed associations. We proposed a framework of potential mechanisms that may contribute to the observed associations. We consider why exposure to *forest outside park* may have a larger effect size than *forest inside park*. We provide explanations for optimal exposure buffer size for significant greenspace types. Last, we discuss the contributions of our findings and identify questions for future research.

### 4.1. Potential mechanisms for observed associations

#### 4.1.1 How might forest and pasture alleviate COVID-19 mortality risk?

We found that forest and pasture exposure were significantly and negatively associated with COVID-19 mortality rates in the US, after control for all covariates. This finding aligns with previous studies (Klompmaker et al., 2021; Lee et al., 2021; Russette et al., 2021; Spotswood et al., 2021). Greenspace may lower COVID-19 mortality risk if it boosts biological processes that fight against the prognosis of COVID-19 (Andersen et al., 2021; Roslund et al., 2020; Roviello et al., 2021), making affected patients less vulnerable to death (Roviello & Roviello, 2021). We suggest that contact with greenspace may reduce mortality rates through increased biogenic volatile organic compound (VOC) exposure, increased environmental microbiota exposure, reduced psychological stress and air pollution, and increased physical activity (Fig. 6).

**Fig. 6.**
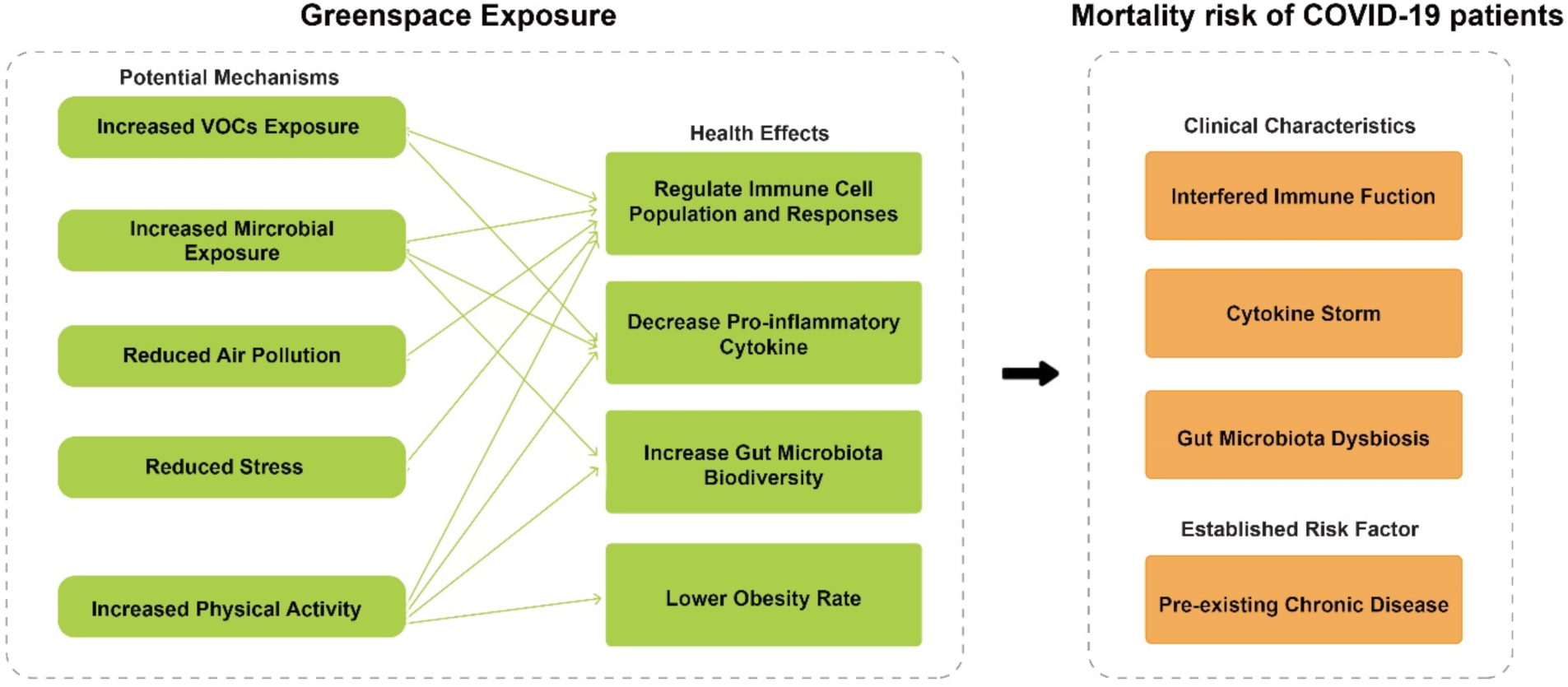
The proposed theoretical model for negative associations between greenspace exposure and COVID-19 mortality rates.

### Exposure to the forest environment increases exposure to biogenic volatile organic compounds (VOCs)

Forests are abundant with phytoncides (e.g., terpenes, limonene and pinene), a group of biogenic VOCs given off by forest trees that have been found to enhance immune capacity and reduce inflammation (Andersen, Corazon, & Stigsdotter, 2021; Cho et al., 2017; Huang et al., 2020; Kim, Song, Cho, & Lee, 2020). Studies suggest forest bathing increases natural killer (NK) cell counts and activity, and the effect can last for more than a month (Li, 2010; Li et al., 2008; Li et al., 2007; Tsao et al., 2018). NK cells activate receptors to recognize virus-infected cells and trigger cytotoxicity (phagocytosis or apoptosis) (Market et al., 2020; Yokoyama, 2005). A recent study found COVID-19 mortality rates are lower in areas with high ratios of hectares of Mediterranean forest per capita, where biogenic VOCs are abundant (Roviello & Roviello, 2021). In addition, studies have shown that nearby greenspaces are associated with a lower level of a biomarker of inflammation - high sensitivity C-reactive protein (Del Valle et al., 2020; Mandel, Harari, Gurevich, & Achiron, 2020; Ribeiro et al., 2019). Forests can increase exposure to VOCs that might boost the NK defenses, compensate for reduced NK cell counts, and modulate excess inflammatory responses in severely affected patients (Market et al., 2020; Osman et al., 2020);

### Exposure to forest and pasture can diversify gut microbiota profile

Though our gut microbial composition is shaped by the interplay of multiple factors, such as diet and genetics (Dhar & Mohanty, 2020), many studies have found microbiome from surrounding green environments can transfer to humans (Grönroos et al., 2019; Parajuli et al., 2018; Parajuli et al., 2020). A 28-day intervention study in Finland found that daily contact with backyard forest and grass areas within 500m can diversify children’s gut microbiota profile and enhance immune capacity (e.g., increases in plasma TGF-β1 levels and the proportion of regulatory T cells) (Roslund et al., 2020). If exposed to greenspaces, the disturbed gut microbe condition and immune function may improve for COVID-19 patients (Claesson et al., 2012; Donati Zeppa, Agostini, Piccoli, Stocchi, & Sestili, 2020; Roslund et al., 2020; Yeoh et al., 2021; Zuo et al., 2020).

### Greenspaces can decrease patients’ exposure to air pollutants

Many studies have noted an inverse association between air pollution and COVID-19 mortality rates (Jaafari, Shabani, Moeinaddini, Danehkar, & Sakieh, 2020; Martelletti & Martelletti, 2020; Shen & Lung, 2017). Chronic exposure to air pollutants was associated with delays in recovery of COVID-19 patients and led to more fatal conditions (Domingo & Rovira, 2020). This may be attributed to modified host respiratory immune responses, perturbed anti-microbial responses, and triggered inflammatory cytokine release from air pollution exposure (Bauer, Diaz-Sanchez, & Jaspers, 2012; Ciencewicki & Jaspers, 2007; Glencross, Ho, Camiña, Hawrylowicz, & Pfeffer, 2020). Forests can reduce air pollutants by intercepting particulate matter on plant surfaces and absorbing gaseous pollutants (Nowak, Hirabayashi, Bodine, & Greenfield, 2014; Nowak, Hirabayashi, Doyle, McGovern, & Pasher, 2018). Trees and forests in the U.S. remove an estimated 17.4 million tonnes (t) of air pollution annually (Nowak et al., 2014). It is thus reasonable to speculate that forests can reduce COVID-19 mortality rates by removing air pollutants.

### Forests can reduce patients’ psychological stress

Patients infected with COVID-19 show a high prevalence of mental problems (Kong et al., 2020; Wang et al., 2021). Mental stress has been linked to dysregulation of the immune system and increased pro-inflammatory cytokines (Gouin et al., 2012; Morey, Boggero et al., 2015; Steptoe et al, 2007). Patients with increased contact with nature or nature views from home during the pandemic were found to have decreased depression and anxiety (Soga et al., 2021). Previous theoretical and empirical studies support nature’s stress-reducing effect (Gidlow et al., 2016; Jiang et al., 2014; Lee et al., 2011; Mancus et al., 2020; Ulrich et al., 1991). It is possible that patients can have strengthened immune function and healthier inflammation level benefit from the stress-reduction effect of contact with forest.

### Green space can promote physical activities (PA) during the pandemic

Since the COVID- 19 outbreak, people have escaped to nature. Recreational activities, such as walking or cycling in parks and trails, have spiked globally during the pandemic (Geng et al., 2021; Lu et al., 2021; Venter et al., 2020; Venter et al, 2021). Physical activity can boost COVID-19 patients’ immune response, modulate inflammation levels, and lower the risk of obesity. Exercise increases NK & T cells, enhances recirculation, and increases lymphocyte concentration and cytotoxic activity (Amatriain-Fernández et al., 2020; Fernandez et al., 2018; Nieman & Wentz, 2019). People who exercise in forested areas can benefit from the synergistic effect from physical activity and exposure to forest (Pretty et al, 2005). Moreover, physical activity has a direct anti-inflammatory effect and can dampen systemic inflammation (Biddle et al., 2019; DeSantis et al., 2012; Nieman & Wentz, 2019). Exercise lowers the risk of obesity, a risk factor for COVID-19. Obesity is the precursor of a range of chronic diseases that have been found to increase COVID-19 mortality (Bastien et al., 2014; Calle & Thun, 2004; Chan et al., 1994; Hussain et al., 2020; Klang et al., 2020; Krauss et al., 1998). Thus, the benefits of greenspace on reduced obesity through physical activity may contribute to the negative association between greenspace and COVID-19 mortality rates (Coombes et al., 2010; De la Fuente et al., 2021; Ghimire et al., 2017; Jia et al., 2020).

#### 4.1.2 How might open space exacerbates COVID-19 mortality risk?

We found the ratio of *open space outside park* is significantly and positively associated with COVID-19 mortality rates. This finding contradicts previous studies that have reported health benefits from exposure to open spaces. Our results suggest that exposure to open space may not be effective or may even be detrimental to COVID-19 mortality rate. Open space in our study is defined as “large-lot single-family housing units or vegetation planted in developed settings” (NLCD, 2016). On one hand, open space can provide health benefits by promoting physical activity, social interaction, and reduced air pollutants (Lu, Chen, et al., 2021), though the effect may not be as strong as it is for forests (Reid, Clougherty, Shmool, & Kubzansky, 2017). On the other hand, the lower supply of open space per capita in urban areas makes it hard to comply with safe social distancing in non-park open spaces (e.g., streets and backyards), and may exacerbate mortality risk. Though outdoor transmission of COVID-19 is rare (Bulfone et al. 2021), people who participate in outdoor social activities such as talking or partying were at higher risk of spreading the disease (Domènech-Montoliu et al. 2021; Peng et al. 2022). Presumably, the higher infection risk might lead to higher COVID-19 mortality rates. More evidence is needed to understand the mechanisms of the mixed effects of open space on COVID-19 mortality rates.

#### 4.1.3 Why might forest outside park have a stronger effect than forest inside park?

We found *forest outside park* to have a larger effect size than *forest inside park* on COVID-19 mortality rates after accounting for other covariates. This finding aligns with previous studies of stronger health-promoting effect outside park areas (Reid et al., 2017; Allard-Poesi et al., 2022). The difference in forest exposure inside and outside park may explain the stronger effect of *forest outside park*. The US population has a ten times greater opportunity to be exposed to *forest inside park* than *forest outside park* within walking distance (Fig. 7). Second, social activities in parks might increase risk of close contact and inhale droplets from infected people (DePhillipo et al., 2021; Praharaj & Han, 2021), thus increase infection risk. Thus, it is reasonable to argue that the increased infection risk caused by the social interactions in parks may offset the other health benefits of *forests inside park* on mortality. Further, the health effect of *forest inside park* may be weakened in part due to shutdown policies in some states, which closed parks due to COVID-19 spread risk (Volenec et al., 2021; Smith et al., 2021).

**Fig. 7.**
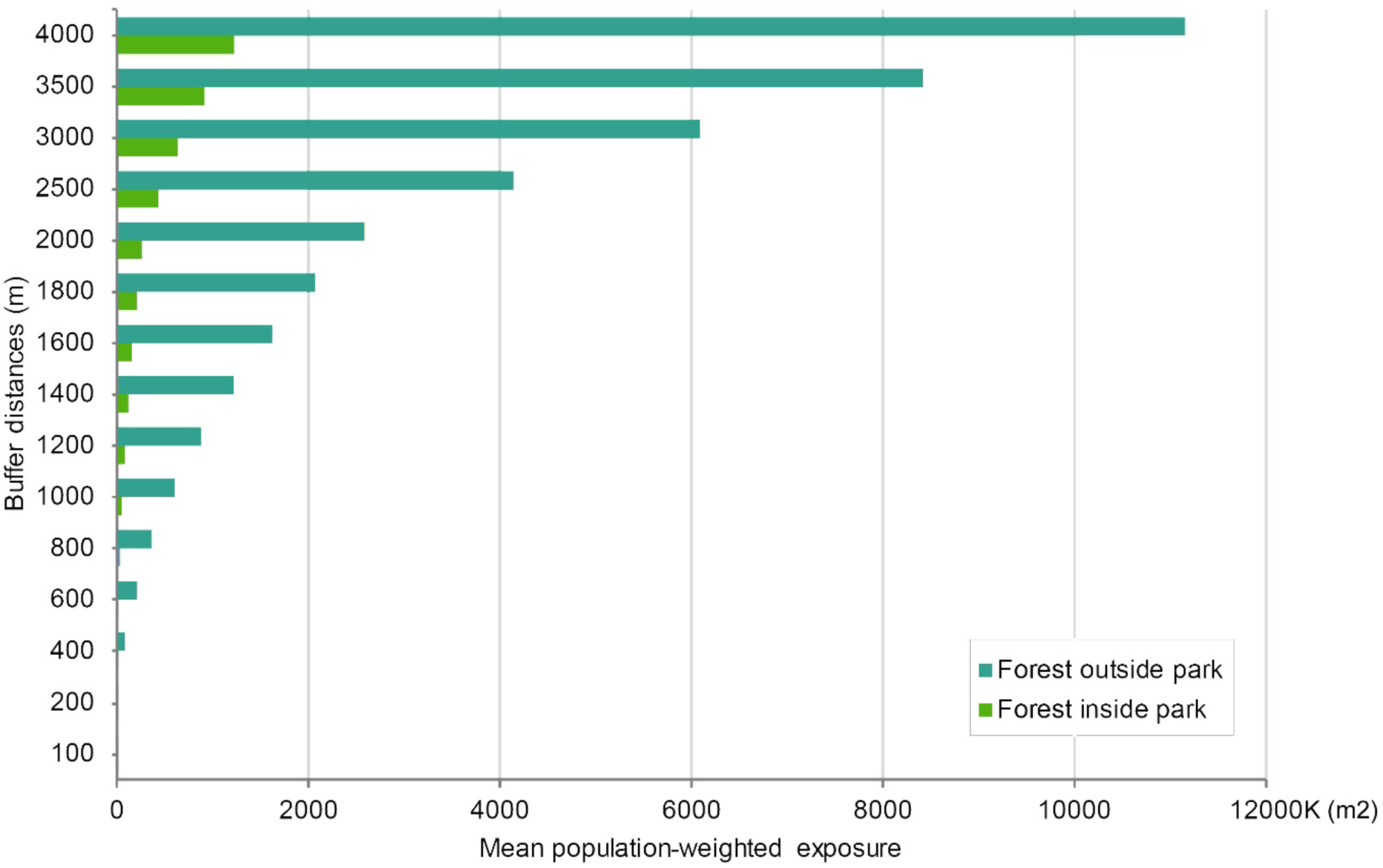
Mean population-weighted exposure to *forest inside park* and *forest outside park* within 4km. The bar represents the average population-weighted forest exposure at county level within each buffer distance.

#### 4.1.4 Optimal exposure buffer distance: what and why?

We found the effect of population-weighted exposure to *forest outside park* increases with a larger buffer distance and reaches an optimal effect at 1km. This suggests that exposure to nearby *forest outside park* within 1km is more effective than exposure to forests that are at a greater walking distance. This may be because nearby forests are visited more often than forests located further away. Studies suggest that the frequency of visits to greenspaces declines as distance increases (Coombes et al., 2010). A distance of 1,120 m (0.7 miles) is the mean walking distance in the U.S. (Yang & Diez-Roux, 2012).

We also found the effect size of *forest inside park* is optimal at 4km, though the effect size is close to that at 400m (2% increase). This suggests the effect of *forest inside park* is less sensitive to buffer size within walking distance. Studies suggest people walk much longer for recreation purposes as opposed to other purposes (Yang & Diez-Roux, 2012). Considering the effect size and previous literature on walking behaviors, we suggest the optimal exposure buffer distance for *forest outside park* to be 1km and the optimal exposure buffer distance for *forest inside park* to be 400m.

### 4.2 Contributions and implications

To our knowledge, this is the first nationwide study distinguish the impact of different types of greenspace on COVID-19 mortality rates in the U.S. In this study, the greenspace exposure measure integrated the dynamics of population distribution and geographic location of greenspaces into exposure assessment, providing a more precise and reasonable exposure estimates. The dose-response associations between different types of green spaces at various buffer distances and COVID-19 mortality rates were examined, allowing us to identify an optimal effect distance, which was previously lacking in the literature.

Evidence from this study suggests that planners and policymakers should prioritize the supply of nearby forests. Specifically, we recommend ensuring that forests outside parks be located within 1000m from residents, and forests inside parks be located within 400m from residents. These forested areas will be especially beneficial in highly urbanized, low-SES, and minority-dominated areas where COVID-19 mortality rates are disproportionally high (Lu et al., 2021). Many greenspaces were temporarily closed during the pandemic to reduce the spread of disease (Ugolini et al., 2020). The findings in this study advocate for keeping nearby greenspace open, especially forested areas. Cities with accessible forested areas can promote health and resilience during the current and future pandemics.

### 4.3 Limitations and future research opportunities

This study has several limitations, which pose opportunities for future research. This is an ecological study using aggregated data at the county level. It is subjected to ecological fallacy. Future studies can use individual level data or experimental studies to confirm the causal relations and the potential underlying mechanisms (Jiang et al., 2021).

Second, the unit of analysis is the county due to the availability of COVID-19 mortality data and other confounding variable. Though county data are widely used in nationwide studies, future studies should use finer-grained data (i.e., census tract level data). Different scales of analyses may reveal different associations between neighborhood greenspace and health outcomes (Richardson et al., 2012).

Third, our research investigated associations using data from 2020, but the situation has continued to evolve with the emergence of vaccines and COVID-19 variants (e.g., Delta and Omicron). Future studies should consider the new situations accordingly.

## 5. Conclusion

Our findings suggest that during 2020, exposure to more green spaces, especially forests, was significantly associated with a lower level of COVID-19 mortality rates, while exposure to more *developed open space* has insignificant or positive associations with the COVID-19 mortality rates. *Forest outside park* is more beneficial than *forest inside park*, with the optimal buffer distance being 1km for *forest outside park*, and within 400m for *forest inside park*. These findings imply that policymakers and planners should prioritize urban greening within optimal distances of residential clusters and keep beneficial greenspaces accessible for COVID-19 and future pandemics.

## Data Availability

All data produced in the present study are available upon reasonable request to the authors

## Acknowledgements

We thank to Long Chen, Xueming Liu, and Xueying Wu for data collection; Bin Chen for methodological help; and Linda Larsen for proof reading the article.

